# Association of Workload and Practice of Respectful Maternity Care Among the Healthcare Providers, Before and During the COVID-19 Pandemic in South Western Nepal: A Cross-Sectional Study

**DOI:** 10.1101/2022.02.21.22271309

**Authors:** Alpha Pokharel, Junko Kiriya, Akira Shibanuma, Ram Silwal, Masamine Jimba

**Affiliations:** Green Tara Nepal, Kathmandu, Nepal; Department of Community and Global Health, Graduate School of Medicine, The University of Tokyo, Tokyo, Japan

## Abstract

**Introduction:** Respectful maternity care is an approach that involves respecting women’s belief, choices, emotions, and dignity during the childbirth process. As the workload among maternity care workforce affects intrapartum quality care, respectful maternity care might have also been affected, particularly during the pandemic. Thus, this study was conducted to examine the association between workload among healthcare providers and their practice of respectful maternity care, before and during the pandemic.

**Methods:** A cross-sectional study was conducted in South Western Nepal. A total of 267 healthcare providers from 78 birthing centers were included. Data collection was done through telephone interviews. The exposure variable was workload among the healthcare providers, and the outcome variable was respectful maternity care practice before and during the COVID-19 pandemic. Multilevel mixed-effect linear regression was used to examine the association.

**Results:** The median client-provider ratio before and during the pandemic was 21.7 and 13.0, respectively. The mean score of respectful maternity care practice was 44.5 (SD 3.8) before the pandemic, which was decreased to 43.6 (SD 4.5) during the pandemic. Client-provider ratio was negatively associated with respectful maternity care practice for both times; before (Coef. −5.16; 95% CI −8.41 to −1.91) and during (Coef. −7.47; 95% CI −12.72 to −2.23) the pandemic.

**Conclusions:** While a higher client-provider was associated with a lower respectful maternity care practice score both before and during the COVID-19 pandemic, the coefficient was larger during the pandemic. Therefore, workload among the healthcare providers should be considered before the implementation of respectful maternity care, and more attention should be given during the pandemic.

## Introduction

Respectful maternity care is an approach based on the principle of ethics and the fundamental rights of women. It includes respecting women’s beliefs, independence, choices, emotions, and dignity while providing maternity care (1). Implementing it in birthing centers reduces unnecessary medical interventions such as episiotomy and fundal pressure. It also improves women’s satisfaction with care and decreases obstetric violence (2). Whereas, disrespectful treatment or negligence during childbirth endangers both mothers’ and newborns’ health and decreases women’s future use of health facilities (2). A large number of women experience disrespect and abuse during childbirth, with the global prevalence varying between 15% and 98% (3). Some common forms of disrespectful maternity care prevalent worldwide are physical abuse, verbal abuse, refusal to provide pain relief, abandonment, poor communication, and lack of privacy (4). As a response to personal safety and security during the COVID-19 pandemic, some of the respectful maternity care practices deteriorated more than before. Restriction of labor companion and breastfeeding were increased during the COVID-19 pandemic (5,6).

An adequate supply of health human resources is essential to delivering respectful maternity care (7). In a qualitative study, healthcare providers reported insufficient staff members, high client loads, and inadequate facilities as hindering factors for better maternity care practice (8). Workload among healthcare providers has caused insufficient and unnecessary client care (2,9). A higher workload among healthcare providers at the birthing centers has decreased interpersonal communication with the clients (10). Increased institutional delivery, but the unreplenished shortage of health human resources has burdened the healthcare providers (11). The inclusion of tasks other than the clinical aspect, such as administrative, monthly reporting, and field works, has further increased their workload (12). Consequently, the tasks that do not directly affect the health of women and newborns are often ignored (2,9). The increased workload has also called upon many malpractices in Low-and Middle-income Countries(LMICs) such as unnecessary use of episiotomy and fundal pressure (10). Moreover, increased workload among healthcare providers has negatively affected the quality of care they provide (13).

Nepal has made substantial progress in improving maternal health care access and utilization (14). Despite this, progress is still required to reach universal access to sexual and reproductive health services by 2030. Along with socio-demographic factors, the low quality of care is also a significant barrier to maternal health care access and utilization in Nepal (15,16). As Nepalese women bypass the primary healthcare centers to have child delivery at the tertiary centers, these centers are overburdened with the clients (17,18). These tertiary health centers were more burdened with clients during the COVID-19 pandemic. The increased work burden during the pandemic further deteriorated the maternal healthcare quality (19).

Respectful maternity care should be understood from both providers’ and mothers’ sides as it is crucial for its effective implementation (20). However, most studies have focused on the mothers’ side, and research on the providers’ side is still lacking (3,8). Since healthcare providers’ scarcity was negatively associated with the quality of skilled birth care (21), it becomes crucial to know if workload among healthcare providers affects the respectful maternity care practice. It is also important to assess the influence of the COVID-19 pandemic on respectful maternity care practice, since the COVID-19 pandemic has further burned the workload among the healthcare providers and negatively affected maternity care (19). Thus, this study was conducted to examine the association of workload among healthcare providers and their respectful maternity care practice, both before and during the COVID-19 pandemic.

## Methods

### Study design and settings

A cross-sectional study was conducted in the South Western Nepal. The study area included four districts: Rupandehi, Kapilvastu, Nawalparasi (Bardaghat Susta West), and Nawalparasi (Bardaghat Susta East). All the government-run birthing centers within the study area were included in the study.

### Participants

All the eligible healthcare providers had a total birthing center experience of more than two years and were working at the present birthing center for more than three months. The duration of work experience was considered to ensure familiarization with the context and work culture (22).

### Variables

#### Exposure variable

The exposure variable was the workload among the healthcare providers. Both subjective and objective measures of workload were assessed. The subjective workload was assessed at individual provider level with the National Aeronautics and Space Administration Task Load Index (NASA TXL) scale (24). It is a scale with a six-item questionnaire that measures the level of mental demand, physical demand, temporal demand, performance, effort, and frustration for the current work (23,24). It consists of a two-part evaluation process: weights and ratings, and its total score range from 0 to 100. A higher score represents a higher workload (24,25).

The objective workload was assessed by the client-provider ratio (26), and the total number of deliveries at the health facility level (10). For the number of deliveries, the healthcare providers were asked to recall the approximate total number of deliveries attained in previous month of data collection. The client-provider ratio was assessed at health facility level and was calculated by dividing the total number of births in the health facility by the total number of healthcare providers during that period.

In Nepal, restrictive measures taken against COVID-19 influenced the client-provider ratio, as the number of institutional births decreased in Nepal (19). To avoid the influence, the client-provider ratio was calculated in two time periods: six months before and after the COVID-19 pandemic declaration by WHO on March 11, 2020 (27). Converting the Nepalese calendar to the Gregorian calendar, the two periods for calculating the client-provider ratio were: July/August 2019 to January/February 2020 (before COVID-19 pandemic declaration) and February/March 2020 to June/July 2020 (after COVID-19 pandemic declaration) (28). To better interpret the client-provider ratio in the regression analysis, the client-provider ratio of six months was converted to per day.

#### Outcome variable

The outcome variable in this study was healthcare providers’ practice of respectful maternity care. It was assessed using a questionnaire adapted from the performance standard for respected maternity care prepared by the Maternal and Child Health Integrated Program (29). The questionnaire is comprised of 7 domain and 27 items: non-abusive care (6 items), consented care (9 items), confidential care (3 items), dignified care (3 items), non-discriminative care (2 items), non-abandonment care (3 items), and non-detention care (1 item) (29). As all the included birthing centers provided free maternity service to the clients, the non-detention domain (detention in a health facility due to inability to pay hospital bills) was removed from the questionnaire (30). Each item has three responses: always (2 points), sometimes (1 point), and never (0 point). The scores of all items were summed to compute the total score. The possible score ranges from 0 to 52, and a higher score represents the better practice of respectful maternity care.

The maternity care practice was also affected by the COVID-19 in Nepal (19). To incorporate the issue, the healthcare providers were asked to report the practice of each item of respectful maternity care before (July/August 2019 to January/February 2020) and during the COVID-19 pandemic (February/March 2020 to June/July 2020) (28).

#### Confounders and covariates

Potential confounders and covariates were added to the study. Healthcare providers’ job positions (7,31) and education level (10,32) were included as confounders. Awareness of respectful maternity care, SBA training, age, being tested positive for COVID-19, and years of job experience were included as covariates (10). Healthcare providers were considered to be aware of respectful maternity care if they had ever heard or read about it in the past.

### Validity and reliability of the scale

Permission was obtained from the concerned organization for the use of the questionnaires. As the questionnaires were not available in the Nepali language, they were checked for cultural adaptation through translation and back translation to the Nepali language and pre-testing of scale among the healthcare providers (33). The respectful maternity care practice questionnaire was not tested for validity and reliability among healthcare providers, and was used in Nepal before. Thus, content validity was ensured through three experts opinion, two online Focus Group Discussions (FGDs) among the healthcare providers, and the pre-test of the questionnaires among the healthcare providers. Cronbach’s alpha was calculated to measure the internal consistency of respectful maternity care practice questionnaire (33). As a whole (26 item), the score was 0.93. Each of the six domains of respectful maternity care scored above 0.80.

### Data collection

Data were collected from September to October 2020. Data were collected through telephone using interviewer-administered questionnaire and response was simultaneously entered into Google forms. The healthcare providers were contacted beforehand for verbal consent and data collection schedule. The research assistants were hired, and trained to obtain verbal consent and to conduct telephone-based interviews. It took approximately 25 minutes to explain the study and interview the healthcare providers. The data for the client-provider ratio were obtained from the records of the health authority.

### Data analysis

Descriptive statistics was performed to present the background characteristics of health facilities and healthcare providers, and practice of respectful maternity care among the healthcare providers. A paired t-test was performed to compare the practice of respectful maternity care before and during the COVID-19 pandemic. A two-level, mixed-effect linear regression analysis was performed with a random intercept at the health facility level. Two null and full models were used for the sub-category of the outcome variable: respectful maternity care practice before and during the COVID-19 pandemic. The intraclass correlation coefficient was used to compare the proportion of variance caused by the random intercept at the health facility level. The significance level was set at 0.05. Google sheet was used for data organization and filtering. Data were exported to R Studio version 1.2.5001 for statistical analyses.

### Research ethics

An ethics approval was obtained from the Research Ethics Committee, Graduate School of Medicine, the University of Tokyo, Japan (serial number: 2020101NI), and Nepal Health Research Council, Nepal (ERB protocol registration number: 524/2020 MT). Permission for data collection was obtained from District Health Offices, district hospitals, and city hospitals. Permission to use healthcare providers’ phone numbers was obtained from them through the head of health facilities. Participation in this study was voluntary. Confidentiality was assured, and verbal consent was taken from each healthcare provider. Documentation of the date and time of consent was done. The audio recording of the telephone interview and the verbal consent were not done.

## Results

### Characteristics of health facility

Out of 79 birthing centers, one province hospital was excluded from the study due to administrative issues. As a result, 78 were included in the study. Kapilvastu district had the highest number of health facilities (n=28), and the Parasi district had the least (n=13). About three-quarters of the health facilities were health posts (Table 1).

**Table 1.** Characteristics of health facilities (n= 78)

| Level of Health Facility | Districts |  |  |  | Total |
| --- | --- | --- | --- | --- | --- |
|  | Kapilvastu | Rupandehi | Nawalpur | Parasi |  |
| Health Post | 23 | 16 | 10 | 8 | 57 |
| PHC<sup>a</sup> | 2 | 5 | 4 | 2 | 13 |
| City Hospital | 2 | 0 | 0 | 2 | 4 |
| District Hospital | 1 | 1 | 1 | 1 | 4 |
| **Total** | **28** | **22** | **15** | **13** | **78** |

<sup>a</sup>PHC: Primary Health Care Center

### Background characteristics of the healthcare providers

A total of 318 healthcare providers were working at the 78 birthing centers. Among them, 267 healthcare providers completed the interview and were included in data analyses. The least number of healthcare provider recruited from a health facility was one, whereas the highest was 11. There were no missing data. Table 2 presents the summary of their background characteristics. The majority of the healthcare providers had undergone Auxiliary Nurse Midwife (ANM) education (70.4%), and SBA training (73.4%), worked as an ANM (88.4%), and worked at a health post (63.3%). About 80% of the healthcare providers had never heard of or read about respectful maternity care in the past, and around 7.0% of them had been tested positive for COVID-19.

**Table 2.**
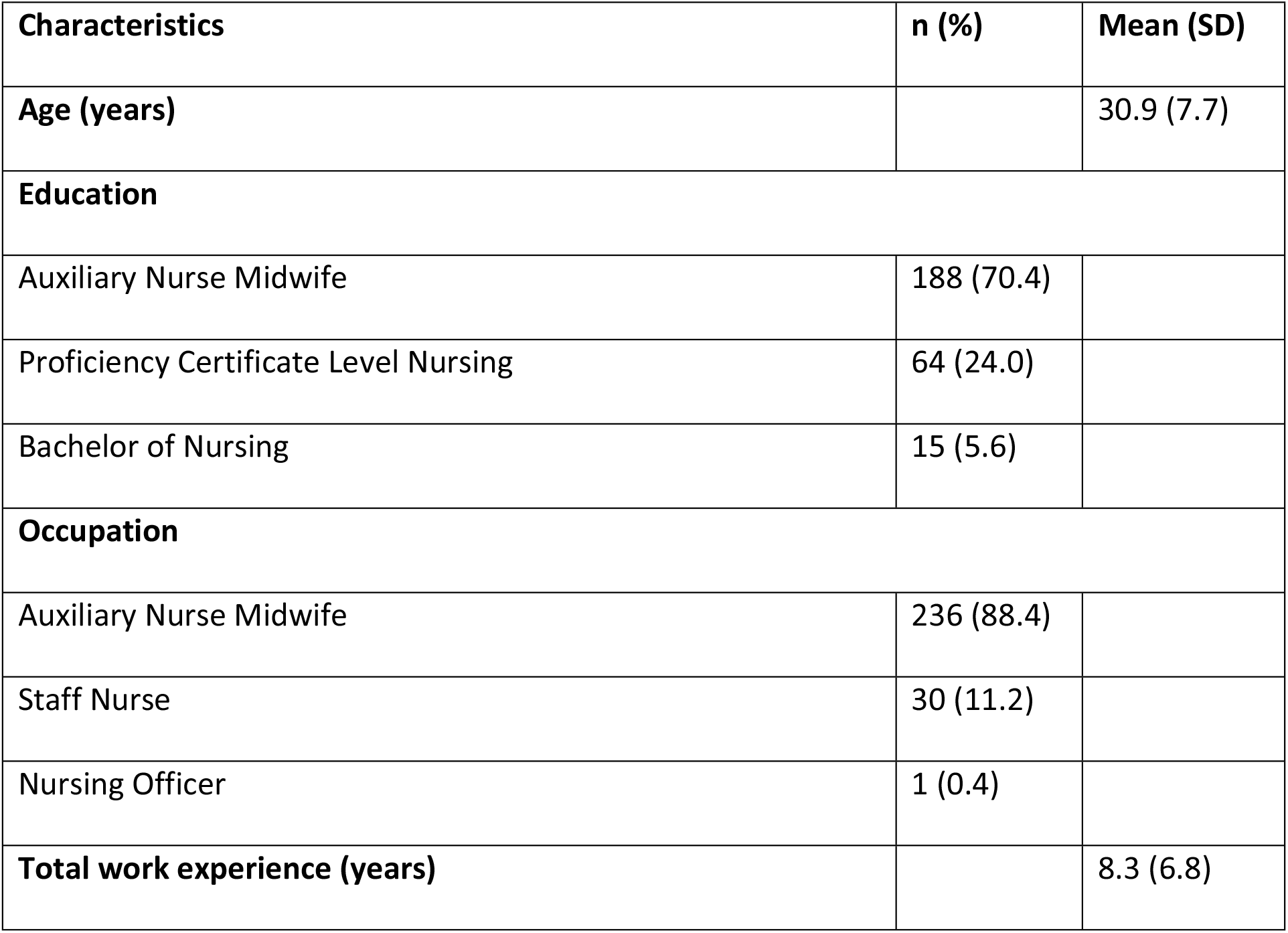
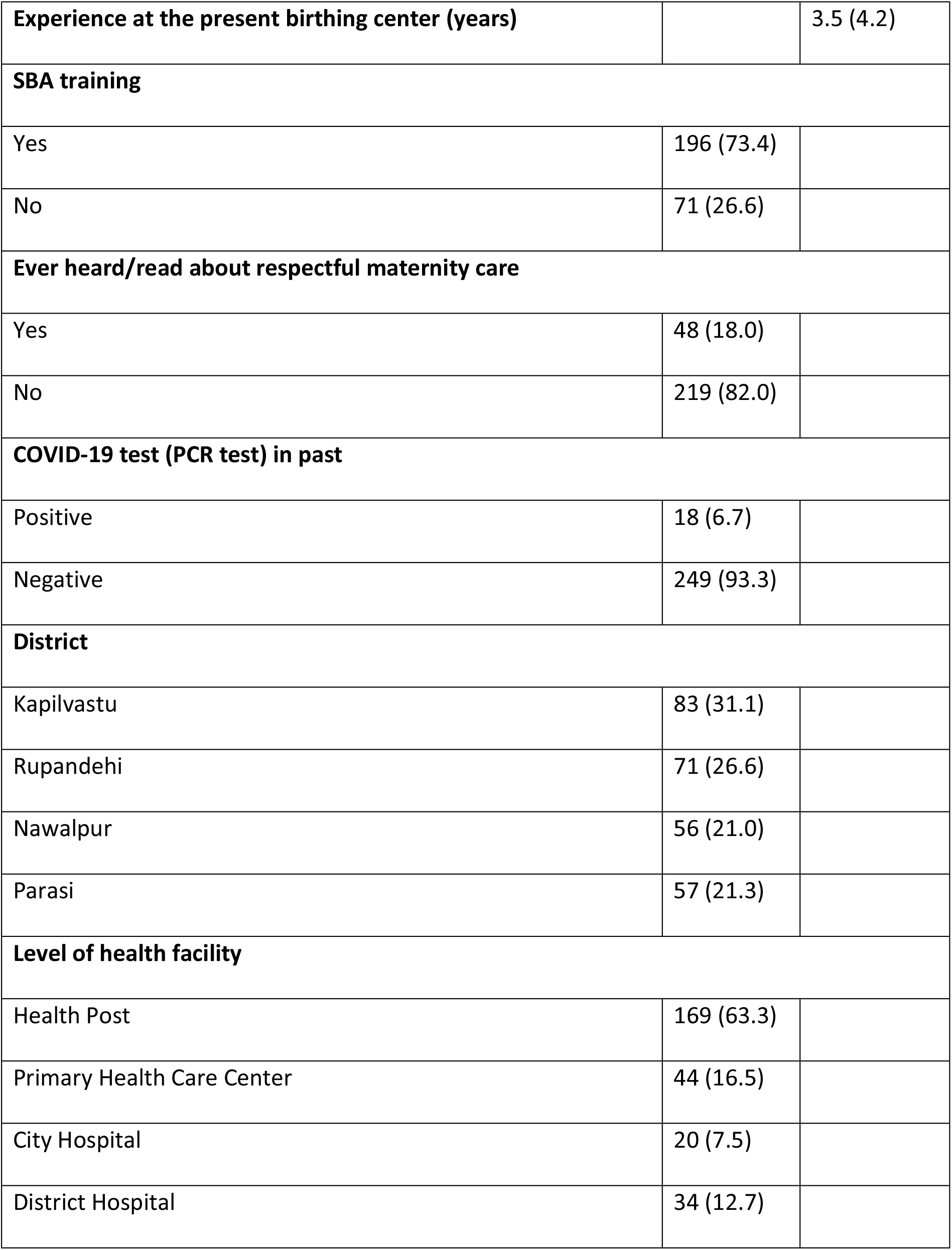
Background characteristics of the healthcare providers (n=267)

| Characteristics | n (%) | Mean (SD) |
| --- | --- | --- |
| **Age (years)** |  | 30.9 (7.7) |
| **Education** |  |  |
| Auxiliary Nurse Midwife | 188 (70.4) |  |
| Proficiency Certificate Level Nursing | 64 (24.0) |  |
| Bachelor of Nursing | 15 (5.6) |  |
| **Occupation** |  |  |
| Auxiliary Nurse Midwife | 236 (88.4) |  |
| Staff Nurse | 30 (11.2) |  |
| Nursing Officer | 1 (0.4) |  |
| **Total work experience (years)** |  | 8.3 (6.8) |
| **Experience at the present birthing center (years)** |  | 3.5 (4.2) |
| **SBA training** |  |  |
| Yes | 196 (73.4) |  |
| No | 71 (26.6) |  |
| **Ever heard/read about respectful maternity care** |  |  |
| Yes | 48 (18.0) |  |
| No | 219 (82.0) |  |
| **COVID-19 test (PCR test) in past** |  |  |
| Positive | 18 (6.7) |  |
| Negative | 249 (93.3) |  |
| **District** |  |  |
| Kapilvastu | 83 (31.1) |  |
| Rupandehi | 71 (26.6) |  |
| Nawalpur | 56 (21.0) |  |
| Parasi | 57 (21.3) |  |
| **Level of health facility** |  |  |
| Health Post | 169 (63.3) |  |
| Primary Health Care Center | 44 (16.5) |  |
| City Hospital | 20 (7.5) |  |
| District Hospital | 34 (12.7) |  |

### Workload among the healthcare providers

The healthcare providers’ mean total subjective workload score was 77.7 (range: 30-100). The median client-provider ratio of six months before and during the COVID-19 pandemic was 21.7 (IQR 9.7 to 52.2) and 13.0 (IQR 6.2 to 28.4). The mean number of deliveries attended by a healthcare provider in the last month was 13.6 (SD 19.6) (Table 3).

**Table 3.** Characteristics of workload among healthcare providers.

| Characteristics | Mean (SD) | Median (1Q-3Q) | Range |
| --- | --- | --- | --- |
| Client-provider ratio of 6 months, before the COVID-19 pandemic (n=263) | 34.9 (34.7) | 21.7 (9.7-52.2) | 0.3-189.0 |
| Client-provider ratio of 6 months, during the COVID-19 pandemic (n=263) | 22.9 (24.0) | 13.0 (6.2-28.4) | 1.2-144.0 |
| Number of deliveries attended in last month (n=267) | 13.6 (19.6) | 9.0 (4.0-15.0) | 0.0-150.0 |
| A score of subjective workload [NASA TXL Scale] (n=267) | 77.7 (13.2) | 78.3 (70.0-87.8) | 30-100 |

### Respectful maternity care practice

During the COVID-19 pandemic, mean total score of respectful maternity care decreased to 43.6 (SD 4.5) from 44.5 (SD 3.8). Table 4 presents the score of practices under each domain of respectful maternity care. For the abuse-free care domain, all healthcare providers never restrained the women during labor. However, 27.3% of them sometimes had to physically or verbally abuse the women. Also, 59% of them always provided pain and comfort relief to the women in labor. The practice of always touching the women in a culturally appropriate way was 95.1 % before the COVID-19 pandemic and 89.5% during the pandemic. All healthcare providers never separated the baby with the mother, both before and during the pandemic.

**Table 4.** Item score of respectful maternity care practice (n=267)

| Questions | Before COVID-19 pandemic |  |  | During COVID-19pandemic |  |  | p-value |
| --- | --- | --- | --- | --- | --- | --- | --- |
|  | Always | Sometimes | Never | Always | Sometimes | Never |  |
|  | (%) | (%) | (%) | (%) | (%) | (%) |  |
| Abuse-free care |  |  |  |  |  |  |  |
| Never physically or verbally abused | 72.2 | 27.3 | 0.0 | 72.0 | 28.0 | 0.0 | 0.318 |
| Never restrained physically | 100.0 | 0.0 | 0.0 | 100.0 | 0.0 | 0.0 | 0.318 |
| Touched in a culturally appropriate way | 95.1 | 4.9 | 0.0 | 89.5 | 10.5 | 0.0 | <0.001 |
| Never separated the baby from mother<sup>a</sup> | 98.5 | 1.5 | 0.0 | 96.2 | 3.8 | 0.0 | 0.057 |
| Never denied food and fluids<sup>a</sup> | 97.0 | 3.0 | 0.0 | 97.0 | 3.0 | 0.0 | 1.000 |
| Provided comfort/pain relief | 58.8 | 23.9 | 17.2 | 59.1 | 17.0 | 23.9 | 0.318 |
| Right to information, informed consent, and choice |  |  |  |  |  |  |  |
| Introduced self to women and her companion | 34.8 | 54.6 | 10.4 | 32.5 | 52.3 | 15.2 | <0.001 |
| Encouraged companion to stay with women | 62.5 | 24.7 | 12.8 | 49.0 | 23.5 | 27.5 | <0.001 |
| Encouraged to ask questions | 91.0 | 9.0 | 0.0 | 85.0 | 14.2 | 0.8 | <0.001 |
| Responded questions promptly and politely | 92.8 | 7.2 | 0.0 | 86.1 | 13.9 | 0.0 | <0.001 |
| Explained the procedure | 90.6 | 9.0 | 0.4 | 84.0 | 14.2 | 1.8 | <0.001 |
| Provided periodic update | 93.2 | 6.8 | 0.0 | 85.7 | 12.7 | 1.6 | <0.001 |
| Allowed to move during labor | 53.5 | 42.6 | 3.7 | 54.0 | 42.3 | 3.7 | 0.318 |
| Allowed to assume the birth position of choice | 19.1 | 32.9 | 48.0 | 19.4 | 32.2 | 48.4 | 0.990 |
| Obtained consent before the procedure | 91.7 | 7.4 | 0.7 | 91.3 | 8.0 | 0.7 | 0.318 |
| **Confidential care** |  |  |  |  |  |  |  |
| Stored file in locked cabinets | 19.1 | 47.9 | 33.0 | 19.0 | 47.7 | 33.3 | 0.157 |
| Used curtain or visual barrier | 56.6 | 34.8 | 8.6 | 56.2 | 38.0 | 7.8 | 0.655 |
| Used drapes and covering | 70.7 | 26.5 | 2.6 | 71.1 | 26.3 | 2.6 | 0.564 |
| **Dignified care** |  |  |  |  |  |  |  |
| Spoke politely to women | 94.0 | 5.6 | 0.4 | 93.0 | 6.6 | 0.4 | 0.083 |
| Allowed non-harmful cultural practice | 95.8 | 4.2 | 0.0 | 95.5 | 4.5 | 0.0 | 0.318 |
| Never insulted and intimidated | 94.3 | 5.7 | 0.0 | 94.3 | 5.7 | 0.0 | 1.000 |
| **Equitable care** |  |  |  |  |  |  |  |
| Spoke in an understandable language | 92.5 | 7.5 | 0.0 | 92.5 | 7.5 | 0.0 | 1.000 |
| Never disrespected & discriminated the women | 98.8 | 1.2 | 0.0 | 98.5 | 1.5 | 0.0 | 0.318 |
| **Non-abandon care** |  |  |  |  |  |  |  |
| Encouraged to call if required | 96.4 | 3.3 | 0.3 | 92.5 | 7.2 | 0.3 | 0.001 |
| Came quickly when called | 67.7 | 32.3 | 0.0 | 64.8 | 35.2 | 0.0 | 0.004 |
| Never left women alone | 56.7 | 41.8 | 0.5 | 54.0 | 42.3 | 3.7 | 0.001 |

<sup>a</sup> unless medically indicated

Regarding the right to information and informed choice care domain, the practice of always allowing a labor companion was 62.5% before the pandemic, and 49.0% during the pandemic. Statistically significant differences in practices for two time points were noticed for: always allowing labor companion, always introducing self to the women and her companion (before and during the pandemic: 34.8% and 32.5%), always encouraging women to ask questions (before and during the pandemic: 91.0% and 85.0%), respond the questions promptly (before and during the pandemic: 92.8% and 86.1%), explain the producers (before and during the pandemic: 90.6% and 84.0%), and provide periodic updates during the COVID-19 pandemic (before and during the pandemic: 93.2% and 85.7%). Allowing women to choose the birth position was never practiced by approximately half of the healthcare providers, both before and during the pandemic.

For the confidential care domain, about one-third of the healthcare providers never stored women’s files in the locked cabinet, both before and during the pandemic. Around half of the healthcare providers sometimes or never used curtains for procedures, both before and during the pandemic.

For non-abandon care domain, 96.4% of the healthcare providers always encouraged the women to call before the COVID-19 pandemic, but it was 92.5% during the pandemic. When called, 67.7% of the healthcare providers always came quickly before the pandemic, but 64.8% came during the pandemic. While the practice of never always leaving the women alone during labor was 56.7% before the pandemic, it was 54.0% during the pandemic.

### Factors associated with respectful maternity care practice

Table 5 presents the results of the multilevel mixed-effect linear regression analysis with a random intercept at the health facility level, for factors associated with respectful maternity care practice by the healthcare providers. The results are presented in two models: respectful maternity care practice before the COVID-19 pandemic and during the pandemic. There are also two null models accompanying each of these full models. Education and total years of experience were excluded from the final model, as they were found to be collinear with occupation and total years of experience, respectively (34).

**Table 5.** Multilevel mixed-effects linear regression analysis for respectful maternity care practice among the healthcare providers.

| Predictors | Respectful Maternity Care Practice |  |  |  |  |  |
| --- | --- | --- | --- | --- | --- | --- |
|  | Before the COVID-19 pandemic |  | P-value | During the COVID-19 pandemic |  | p-value |
|  | Null model | Final Model (n=267) |  | Null model | Final Model (n=267) |  |
|  |  | Estimates (CI) |  |  | Estimates (CI) |  |
| Healthcare provider characteristics (Fixed effect) |  |  |  |  |  |  |
| The subjective measure of workload |  | 0.02 (-0.02 to 0.05) | 0.376 |  | 0.02 (-0.02 to 0.06) | 0.246 |
| Experience at the present birthing center |  | -0.04 (-0.15 to 0.07) | 0.450 |  | -0.02 (-0.14 to 0.11) | 0.784 |
| Occupation |  |  |  |  |  |  |
| Auxiliary Nurse Midwife |  | Reference |  |  | Reference |  |
| Staff Nurse |  | -0.12 (-1.62 to 1.39) | 0.879 |  | -0.004 (-1.68 to 1.69) | 0.996 |
| Age |  | 0.08 (0.02 to 0.14) | 0.011 |  | 0.07 (0.002 to 0.14) | 0.037 |
| Ever heard/read about RMC |  |  |  |  |  |  |
| No |  | Reference |  |  | Reference |  |
| Yes |  | 0.68 (-0.37 to 1.73) | 0.203 |  | 1.38 (0.17 to 2.59) | 0.055 |
| **Skilled Birth Attendant training** |  |  |  |  |  |  |
| No |  | Reference |  |  | Reference |  |
| Yes |  | -0.20 (-1.15 to 0.75) | 0.684 |  | -0.36 (-1.43 to 0.71) | 0.513 |
| **COVID-19 test (PCR test) in past** |  |  |  |  |  |  |
| Negative/ Not tested |  | - |  |  | Reference |  |
| Positive |  | - |  |  | -3.18 (-5.06 to -1.30) | 0.001 |
| **Number of deliveries attended last month** |  | -0.02 (-0.04 to 0.01) | 0.129 |  | -0.02 (-0.04 to 0.01) | 0.236 |
| **Health facility level characteristics (n=78)** |  |  |  |  |  |  |
| **Client-provider ratio per day** |  |  |  |  |  |  |
| Before the COVID-19 pandemic* |  | -5.16 (-8.41 to -1.91) | 0.002 |  |  |  |
| During the COVID-19 pandemic |  |  |  |  | -7.47 (-12.72 to -2.23) | 0.005 |
| **Level of health facility** |  |  |  |  |  |  |
| Health Post |  | 0.48 (-1.39 to 2.36) | 0.614 |  | -0.12 (-2.41 to 2.18) | 0.921 |
| Primary Health Care Center |  | -0.55 (-2.69 to 1.58) | 0.611 |  | -1.93 (-4.57 to 0.71) | 0.151 |
| Hospital (City + District) |  | Reference |  |  | Reference |  |
| **Random effect** |  |  |  |  |  |  |
| **Health facility level variance (SD)** | 5.04<br>(2.24) | 3.25 (1.80) |  | 8.36<br>(2.89) | 5.60 (2.36) |  |
| **ICC (%)** | 34.0 | 26.0 |  | 40.0 | 33.0 |  |

CI: Confidence Interval, SD: Standard Deviation, ICC: Intraclass Correlation Coefficient, Before the COVID-19 pandemic: July/August
2019 to January/February 2020, During the COVID-19 pandemic: February/March 2020 to June/July 2020.

The healthcare providers’ characteristics were considered as level 1 variable, and health facility characteristics were considered as level 2 variable. The health facilities in the null model explained 34.0% variance for the respectful maternity care practice before the COVID-19 pandemic. After controlling for exposure variables, 26% of the variation was explained by the health facilities. The null model explained 40% variance of practice among the health facilities, during the COVID-19 pandemic. After controlling the exposure variables, it was 33.0%.

Age of the healthcare providers was positively associated with respectful maternity care practice before (Coef. 0.08; 95% CI 0.02 to 0.14) and during (Coef. 0.07; 95% CI −0.002 to 0.14) the COVID-19 pandemic. Being tested positive for COVID-19 in the past was negatively associated with respectful maternity care practice during the COVID-19 pandemic (Coef. −3.18; 95% CI −5.06 to −1.30). The client-provider ratio per day was negatively associated with respectful maternity care practice before (Coef. − 5.16; 95% CI −8.41 to −1.91) and during (Coef. −7.47; 95% CI −12.72 to −2.23) the COVID-19 pandemic.

## Discussions

### Principal findings

This study investigated the association between workload and practice of respectful maternity care among the healthcare providers in South Western Nepal, before and during the COVID-19 pandemic. While the workload among the healthcare providers was negatively associated with respectful maternity care practice, both before and during the pandemic, the effect of workload on the practice of respectful maternity care was larger during the pandemic.

Higher workload among the healthcare providers was negatively associated with respectful maternity care practice. However, the coefficient was larger during the pandemic (Coef. −7.47; 95% CI −12.72 to −2.22) than before (Coef. −5.16; 95% CI −8.41 to − 1.91). In a high workload setting, healthcare providers often ignore the procedures that do not immediately impact the mother and baby’s life (35,36). These high workload settings are often located in urban areas, where the COVID-19 cases are high (5,36). Since healthcare providers want to prevent themselves from acquiring COVID-19 infection, they avoid respectful maternity care practices such as client communication and labor companionship (5). As, a result the effect of workload on respectful maternity care practice might have been greater during the pandemic in this study. A study from Nepal also found decrease in labor companionship during the pandemic (19). Therefore, decrement of workload among the healthcare providers should be considered more during the pandemic.

The variance among each health facility for respectful maternity care practice increased more during the pandemic. The difference in client-provider ratio found among the health facilities from this study, could be a possible reason for the variance of respectful maternity care across the health facilities. It was also found from the national data that the decline in delivery cases varied across the type of health facility in Nepal (28). While 50% or more of local health facilities were closed during the early lockdown period for child delivery, almost all tertiary healthcare centers were open (28). Strengthening the quality of local health facilities can increase child delivery cases there, which will help distribute client-provider ratio among health facilities (37). As a result, respectful maternity care can be practiced, even during the pandemic.

Increased age among the healthcare providers was positively associated with the respectful maternity care practice. A possible explanation could be, with age healthcare providers become more experienced with maternity care. They become more skillful in satisfying women’s need, and respecting her desires (10).

Healthcare providers who were tested positive for COVID-19 had lower respectful maternity care practice scores. Healthcare providers are praised for their work during the COVID-19 pandemic. However, they are still stigmatized by community for being the source of infection (38,39). To avoid being stigmatized, they might have avoided respectful maternity care practices such as client interaction and communication (39,40). Also, other possible reasons could be decreased mental and physical performance due to stigma, psychological distress, and diseases (41,42). Therefore, more physical and mental health support to the healthcare providers should be provided to improve their performance (43).

### Strengths and weaknesses

This study has several strengths. It is one of the first on healthcare providers’ workload and its association with respectful maternity care during the COVID-19 pandemic. As this study included all the health facilities in the study area, it provides an overview of all levels of health facilities and all the cadre of healthcare providers in Nepal.

It also has several limitations. The respectful maternity care practice was affected by social desirability bias. In order to overcome it, assurance of anonymity and confidentiality was provided. Also, the healthcare providers were well explained about the objectives of the study and its possible impact on the scientific literatures. The data related to respectful maternity care practice before the pandemic depended on the healthcare provider’s memory, which might have introduced a measurement error in the outcome assessment. Since the COVID-19 pandemic occurred during the data collection, the researcher had to rely on the self-reporting data for before the pandemic period. The visual rating of the NASA TXL scale was changed to auditory description, as the data collection was done through telephone for the prevention of spread of COVID-19 infection. The change of scale from visual to auditory could have caused over-reporting or under-reporting of the workload. This could be the reason why it did not show strong evidence for the association with the respectful maternity care practice (44).

## Conclusions

While a higher client-provider ratio was associated with a lower respectful maternity care practice score both before and during the COVID-19 pandemic, the coefficient was larger during the pandemic. Also, the variance among the health facilities for respectful maternity care was increased more during the COVID-19 pandemic. The findings of this study call for the decrement in the client-provider ratio for better respectful maternity care practice, especially during the pandemic. However, supplying health human resources per the population demand, especially during the pandemic, may be difficult in a resource-limited setting. The number of delivery at local health facilities can be increased by improving the quality of care, particularly during the pandemic. This could help equal distribution of client-provider and practice respectful maternity care, even during the pandemic. Further, the healthcare providers who were tested positive for COVID-19 had lower respectful maternity care practice scores. Therefore, additional physical and mental health support to the healthcare providers should be considered to improve their respectful maternity care practice, particularly during the pandemic.

## Data Availability

The dataset will be uploaded in public repository, and a link/DOI will be provided. The dataset has not been uploaded now but we will do it after the article gets accepted. However, if required, it can be done prior to acceptance as well.

## Acknowledgments

The authors would like to express sincere gratitude to all healthcare providers who participated in this study. We want to thank the District Public Health Office of Rupandehi, Kapilvastu, Nawalparasi (East and West), and Green Tara Nepal for assisting our research. We are thankful to all health facilities’ administrative departments for supporting and facilitating our research. We are grateful to Professor Nirmala Pokharel, Ms. Yasoda Giri, and Ms. Namita Shrestha for their valuable and expert contribution to the questionnaire’s content validity. Special mention of thanks to Ms. Kopila Shrestha and Mr. Kumar Thapa for helping with the administrative process. We are thankful to Ms. Bandana Rajbanshi and Ms. Renu Chaudhary for helping with the pre-test of the questionnaire. We are grateful to Ms. Asmita Jha, Engila Khadka, and Avida Ojha for helping with data collection. We would also like to thank Ms. Suhyoon Choi for helping with the documentation process.

## References

1. The White Ribbon Alliance for Safe Motherhood. Respectful maternity care: The universal rights of childbearing women. The White Ribbon Alliance for Safe Motherhood; 2011.

2. World Health Organization. Intrapartum care for a positive childbirth experience. Geneva: World Health Organization; 2018.

3. Afulani PA, Moyer CA. Accountability for respectful maternity care. Lancet. 2019; 394(10210):1692–3.

4. Bohren MA, Vogel JP, Hunter EC, Lutsiv O, Makh SK, Souza JP, et al. The mistreatment of women during childbirth in health facilities globally: A mixed-methods systematic review. PLoS Med. 2015; 12(6):1–32.

5. Jolivet RR, Warren CE, Sripad P, Ateva E, Gausman J, Mitchell K, et al. Upholding rights under COVID-19: The respectful maternity care charter. Health Hum Rights. 2020; 22(1):391–4.

6. Rocca-Ihenacho L, Alonso C. Where do women birth during a pandemic? Changing perspectives on safe motherhood during the COVID-19 pandemic. J Glob Heal Sci. 2020;2(1):4.

7. Bowser D, Hill K. Exploring evidence for disrespect and abuse in facility-based childbirth report of a landscape analysis. Boston: Harvard School of Public Health University Research Co., LLC;2010.

8. Ndwiga C, Warren CE, Ritter J, Sripad P, Abuya T. Exploring provider perspectives on respectful maternity care in Kenya: “work with what you have.” Reprod Health. 2017;14(1):99.

9. Bogren M, Erlandsson K, Akter HA, Khatoon Z, Chakma S, Chakma K, et al. What prevents midwifery quality care in Bangladesh? A focus group enquiry with midwifery students. BMC Health Serv Res. 2018;18(1):639.

10. Dynes MM, Twentyman E, Kelly L, Maro G, Msuya AA, Dominico S, et al. Patient and provider determinants for receipt of three dimensions of respectful maternity care in Kigoma Region, Tanzania-April-July, 2016. Reprod Health. 2018;15(1):41 The White Ribbon Alliance. Respectful maternity care. Vol. 26. 2016.

11. World Health Organization. Global strategy on human resources for health: Workforce 2030. Geneva: World Health Organization; 2016.

12. Global health workforce alliance. Country responses [Internet]. global health workforce alliances; 2020 [cited 2020 Dec 3]. Available from: https://www.who.int/workforcealliance/countries/npl/en/.

13. Chang LY, Yu HH, Chao YFC. The Relationship Between Nursing Workload, Quality of Care, and Nursing Payment in Intensive Care Units. J Nurs Res. 2019;27(1):1.

14. Ministry of Health, New ERA. Nepal Demographic Health Survey NDHS 2016 final report. Singha Durbar, Nepal: Ministry of Health, New ERA; 2017.

15. Kumar Aryal K, Sharma SK, Nath Khanal M, Bista B, Lal Sharma S, Kafle S, et al. Maternal health care in Nepal: trends and determinants. DHS Further Analysis Reports No. 118. Rockville, Maryland, USA; 2018.

16. Shah R, Rehfuess EA, Paudel D, Maskey MK, Delius M. Barriers and facilitators to institutional delivery in rural areas of Chitwan district, Nepal: A qualitative study. Reprod Health. 2018;15(1):110.

17. Roder-Dewan S, Nimako K, Twum-Danso NAY, Amatya A, Langer A, Kruk M. Health system redesign for maternal and newborn survival: Rethinking care models to close the global equity gap. BMJ Global Health. 2020;5(10):2539.

18. FHD/NHSSP. Responding to increased demand for institutional childbirths at referral hospitals in Nepal: situational analysis and emerging options, 2013.

19. Kc A, Gurung R, Kinney M V, Sunny AK, Moinuddin M, Basnet O, et al. Effect of the COVID-19 pandemic response on intrapartum care, stillbirth, and neonatal mortality outcomes in Nepal: a prospective observational study. Lancet Glob Health. 2020;8(10):e1273–e1281.

20. Jolly Y, Aminu M, Mgawadere F, Van Den Broek N. We are the ones who should make the decision - knowledge and understanding of the rights-based approach to maternity care among women and healthcare providers. BMC Pregnancy Childbirth. 2019;19(1) 42.

21. Onta S, Choulagai B, Shrestha B, Subedi N, Bhandari GP, Krettek A. Perceptions of users and providers on barriers to utilizing skilled birth care in mid-and far-western Nepal: a qualitative study. Glob Health Action. 2014;7(1):24580. Dec 2];7(1):24580.

22. Asefa A, Bekele D, Morgan A, Kermode M. Service providers’ experiences of disrespectful and abusive behavior towards women during facility based childbirth in Addis Ababa, Ethiopia. Reprod Health. 2018;15(1):4.

23. Hart SG, Staveland LE. Development of NASA-TLX (Task Load Index): Results of empirical and theoretical research. Advances in Psychology. 1988;52(C):139–83.

24. Human Performance Research Group, NASA Ames Research Center. TASK LOAD INDEX (NASA-TLX) v 1.o.Moffett Field. California: NASA Ames Research Center.

25. Hoonakker P, Carayon P, Gurses AP, Brown R, Khunlertkit A, McGuire K, et al. Measuring workload of ICU nurses with a questionnaire survey: the NASA Task Load Index (TLX). IIE Trans Healthc Syst Eng. 2011;1(2):131–43.

26. Okonofua F, Ntoimo L, Ogu R, Galadanci H, Abdus-Salam R, Gana M, et al. Association of the client-provider ratio with the risk of maternal mortality in referral hospitals: A multi-site study in Nigeria. Reprod Health. 2018;15(1):1–9.

27. World Health Organization. WHO Director-General’s opening remarks at the media briefing on COVID-19 - 11 March 2020 [Internet]. Geneva: World Health Organization; 2020 [cited 2021 Jan 4]. Available from: https://www.who.int/director-general/speeches/detail/who-director-general-s-opening-remarks-at-the-media-briefing-on-covid-19---11-march-2020

28. Government of Nepal, Ministry of Health and Population, Department of Health Service, Management Division, Integrated Health Information Management Section. Assess impact of COVID-19 pandemic in selected health services with estimation of ‘ excess maternal deaths. Kathmandu, Nepal: Ministry of Health, Government of Nepal, Ministry of Health and Population, Department of Health Service, Management Division, Integrated Health Information Management Section; 2021.

29. USAID, MCHIP. espectful maternity care standards. Washington DC, United States: MCHIP, USAID. 2019.

30. Government of Nepal, Ministry of Health and Population. Safe Motherhood Programme. [Internet]. Government of Nepal, Ministry of Health and Population; 2020 [cited 2020 Dec 4]. Available from: https://www.mohp.gov.np/eng/program/reproductive-maternal-health/safe-motherhood-programme.

31. Friese MA, White S V., Byers JF. Chapter 34. Handoffs: Implications for Nurses. Patient Saftey and Quality: An evidence-based handbook for nurses. Rockville (MD): Agency for healthcare research and quality (US); 2008. Chapter 34.

32. Gebeyehu S, Zeleke B. Workplace stress and associated factors among healthcare professionals working in public health care facilities in Bahir Dar City, Northwest Ethiopia, 2017. BMC Res Notes. 2019; 12(1): 1–5.

33. Streiner DL, Norman GR. Health Measurement Scales: A practical guide to their development and use. Health Measurement Scales: A Practical guide to their development and use. Fourth edition. Newyork: Oxford University Press; 2008.

34. Dormann CF, Elith J, Bacher S, Buchmann C, Carl G, Carré G, et al. Collinearity: a review of methods to deal with it and a simulation study evaluating their performance. Ecography (Cop). 2013 Jan; 36(1):27–46.

35. Reader TW, Gillespie A. Patient neglect in healthcare institutions: A systematic review and conceptual model. BMC Health Serv Res. 2013;13:156.

36. Kafle K, Shrestha DB, Baniya A, Lamichhane S, Shahi M, Gurung B, et al. Psychological distress among health service providers during COVID-19 pandemic in Nepal. PLoS One. 2021;16(2):e0246784.

37. Kimani RW, Maina R, Shumba C, Shaibu S. Maternal and newborn care during the COVID-19 pandemic in Kenya: Re-contextualising the community midwifery model. Hum Resour Health. 2020;18(1):75.

38. Taylor S, Landry CA, Rachor GS, Paluszek MM, Asmundson GJG. Fear and avoidance of healthcare workers: An important, under-recognized form of stigmatization during the COVID-19 pandemic. J Anxiety Disord. 2020;75:102289.

39. Khanal P, Devkota N, Dahal M, Paudel K, Joshi D. Mental health impacts among health workers during COVID-19 in a low resource setting: a cross-sectional survey from Nepal. Global Health. 2020; 16(1):89.

40. Razu SR, Yasmin T, Arif TB, Islam MS, Islam SMS, Gesesew HA, et al. Challenges Faced by Healthcare Professionals During the COVID-19 Pandemic: A Qualitative Inquiry From Bangladesh. Front Public Heal. 2021; 9:1024.

41. Suvvari TK, Kutikuppala LVS, Tsagkaris C, Corriero AC, Kandi V. Post-COVID-19 complications: Multisystemic approach. Journal of Medical Virology. 2021; 93(12):6451–5.

42. Ramaci T, Barattucci M, Ledda C, Rapisarda V. Social Stigma during COVID-19 and its Impact on HCWs Outcomes. Sustainability. 2020; 12(9):3834.

43. Liu Q, Luo D, Haase JE, Guo Q, Wang XQ, Liu S, et al. The experiences of health-care providers during the COVID-19 crisis in China: a qualitative study. Lancet Glob Health. 2020;8(6):e790–8.

44. Block ES, Erskine L. Interviewing by telephone: Specific considerations, opportunities, and challenges. Int J Qual Methods. 2012;11(4):428–45.

